# Ultrasound-detected accelerated fat deposition in the vastus medialis as a marker for early knee osteoarthritis

**DOI:** 10.1101/2023.04.16.23288640

**Authors:** Atsushi Hoki, Ella D’Amico, Fabrisia Ambrosio, Yoshikazu Matsuda, Hirotaka Iijima

## Abstract

**Importance:** The absence of a reliable marker of early-stage knee osteoarthritis (KOA) represents a missed opportunity to develop interventions that restore joint integrity. The use of ultrasonography (US) to measure fat infiltration of muscle, which has been associated with incidence and progression of KOA, is one possible solution. Summarizing the trajectory of fatty infiltration during natural aging and KOA processes and establishing US-implementation as a tool for the early diagnosis of KOA represent essential first steps.

**Objective:** (1) To summarize the alterations in quadriceps muscle quality during healthy aging (young vs. older) and in the setting of KOA (older vs. KOA) using computational approaches, and (2) to validate and translate these findings by applying US assessment of muscle quality in a clinical setting.

**Design, Setting, Participants:** Systematic review with network meta-analysis integrated with a subsequent case-control validation study in an orthopedic clinic. Both analyses evaluated three groups: healthy young, healthy older, and individuals with KOA.

**Main outcome and Measures:** Quadriceps muscle quality assessed by US.

**Results:** Data from a total of 718 healthy aging individuals (young, n = 336; old, n = 382) and 1,046 individuals with KOA (old, n = 625; KOA, n = 421) were synthesized for meta-analyses. As determined by US, older adults displayed poor quadriceps muscle quality compared to young; this decline was significantly exacerbated when individuals had KOA. The network meta-analysis revealed that the presence of KOA significantly accelerated muscle fatty infiltration compared to natural aging. Notably, US findings were comparable to gold-standard imaging modalities of both MRI and CT for fat infiltration in skeletal muscle. The findings from these computational analyses were supported by an US-implemented validation study in a clinical setting, which also identified the vastus medialis as the most sensitive quadricep muscle to predict both the onset and development of KOA.

**Conclusions and Relevance:** This study established accelerated fat deposition in the vastus medialis as a distinct marker for early KOA, as reliably identified by US. These findings expand opportunities for the early diagnosis and treatment for KOA in a clinical setting given the increased cost-efficiency and accessibility of the US system compared to MRI or CT.

**Trial registration:** PROSPERO Identifier: CRD 42022380856

**KEY POINTS:** *Question:* Does ultrasonography identify distinct declines in quadriceps muscle quality related to age and knee osteoarthritis (KOA)?

*Findings:* A hybrid approach of network meta-analysis (1,794 participants) followed by ultrasound assessment revealed that KOA significantly accelerated aging-induced fat infiltration in muscle. This study suggests that the 10-year rate of decline in quadriceps muscle quality in KOA is greater than in healthy aging. Furthermore, these changes can sensitively be assessed by ultrasonography.

*Meaning:* Fatty infiltration in the quadriceps muscle over the course of natural aging is accelerated in the setting of KOA. Such trajectories are assessable using ultrasonography, a valuable approach to identify adults at risk for KOA.

## INTRODUCTION

As individuals are living longer than ever before, an enhanced understanding of how to effectively promote healthy aging and prevent age-related diseases in older adults becomes increasingly important. One of the most pervasive age-related diseases is knee osteoarthritis (KOA).^1^ Between 1990 to 2019, the number of years lived with KOA-induced disability increased by 39%,^2^ and in 2019, KOA represented the 15th leading cause of disability worldwide.^2^ Escalating clinical interest is focused on the development of accurate diagnosis and efficacious intervention early in the disease and prior to the onset of major structural damage and debilitating symptoms.^3^

One candidate for disease screening is quantification of early-stage alterations in quadriceps muscle composition. In 2022, Øiestad *et al* published a systematic review that included a meta-analysis of longitudinal studies concluding that quadriceps muscle weakness increased the odds of KOA incidence.^4^ While quadriceps muscle weakness in individuals with KOA is often attributed to decreased to muscle mass, muscle quality also plays a critical physiologic role in force production and metabolism.^5^ Accumulated fat in the muscle secrets adipokines, which are thought to mediate adipose-cartilage crosstalk and ultimately contribute to the pathogenesis of KOA.^6,7^ A recent animal study showed that the knee joints of lipodystrophic mice, which lack adipose tissue and, thus, adipokine signaling, were protected from spontaneous or posttraumatic KOA, despite having body weights and systemic inflammatory levels similar to high fat mice.^8^ Supporting the role of the adipose-cartilage axis in the development of KOA, previous studies have shown that the accumulation of non-contractile (i.e., adipose) tissue in quadriceps muscle is increased in individuals with KOA^9^ and is associated with KOA progression, independent of a change in muscle mass and strength.^10^

Muscle quality is reliably quantified by the studies that assess the correlation between intramuscular adipose tissue and radiographic and/or symptomatic KOA using magnetic resonance imaging (MRI) and computed tomography (CT),^9^ which are two gold standard methods to assess muscle fat infiltration at a research stage. If signs of progression are accurately detected at an earlier stage of KOA, interventions such as therapeutic exercise and self-management (e.g., educational intervention for coping and self-efficacy skills)^11^ can be more effective in preventing and/or delaying KOA progression.^12^ However, these imaging modalities are not typically used for screening of adults at risk of KOA in clinical practice owing to their financial and time burden.^13,14^

To overcome the barriers of MRI and CT, the use of diagnostic ultrasound, or ultrasonography (US), is increasingly appreciated as a potential alternative to detect subtle changes in muscle quality seen with early KOA. For the assessment of muscle quality, US is an accessible and low cost tool compared to MRI and CT, and reliable equivalent to these gold standard methods,^15,16,17^ that would allow for bedside diagnosis. A comprehensive systematic review of US characterizing the alterations in muscle quality during natural aging compared to KOA is a logical first step to address this existing gap and establish US as a tool to diagnose early KOA.

Here, we thoroughly summarized the existing evidence on quadriceps muscle quality in the context of aging and KOA as assessed by US. We also compared our findings to those observed by MRI and CT. First, we assessed differences in quadriceps muscle quality between healthy young and older adults to define changes seen with natural aging. Next, we characterized pathology-related changes in quadriceps muscle quality by comparing people with KOA to healthy age-matched adults. Using a network meta-analysis approach, we summarized the decline in quadriceps muscle quality associated with healthy aging (i.e., healthy young vs healthy older adults) and the pathological KOA process (i.e., healthy young vs individuals with KOA) to delineate the trajectory of quadriceps muscle quality decline within each cohort. Finally, the findings from these computational analyses were cross-checked by a case-control study comparing differences in muscle quality evaluated by US between healthy young, healthy older and individuals with KOA. This proof-of-concept trial also sought to identify differences by target muscle and by KOA severity. We anticipate these findings can be used to establish US assessment of muscle quality as a means of identifying individuals at high risk of KOA in the clinical setting.

## RESULTS

A systematic search summarizing age- and KOA-related decline in muscle quality identified a total of 287 articles from electronic databases, of which 22 met our inclusion criteria (14 studies for young vs. older [718 participants]; 8 studies for older vs. KOA [1,046 participants]; **Figure S1**). **Table 1** summarizes characteristics of the included studies. Individuals with KOA displayed mild to severe disease (KL grade 1-4) and mild to moderate clinical symptoms (**Table S1**). **Table S2** shows a summary of measurement methods. Five out of eight (62.5%) MRI studies used intramuscular fat volume as an outcome, as calculated by chemical shift-based water-fat separation method known as the Dixon technique.^18^ Two thirds (66.7%) of CT studies adopted Hounsfield Unit (HU), a marker of intramuscular fat content.^19^ All studies with US adopted echo intensity as an outcome, which represents intramuscular adipose and fibrous tissue^20^. Echo intensity was additionally modified in some studies for the attenuation of ultrasound waves due to subcutaneous fat tissue.^15^ Intra- and inter-rater reliability (intra-rater or inter-rater) of US was good to excellent but lower than CT and MRI due to a wide confidence interval (**Table S2**). **Table S3** summarizes the main outcomes. Notably, target muscles of US assessments showed high inter-trial variability. The majority of studies targeted either the vastus medialis (VM) or a combination of two or more individual quadriceps muscles (RF, VM, vastus lateralis [VL], or vastus intermedius [VI]). However, a few studies targeted the rectus femoris (RF) muscle only. Most of the studies in young vs. older individuals used US (9 [64.3%] studies) and those in older vs. KOA used MRI (4 [50.0%] studies).

**Table 1.**
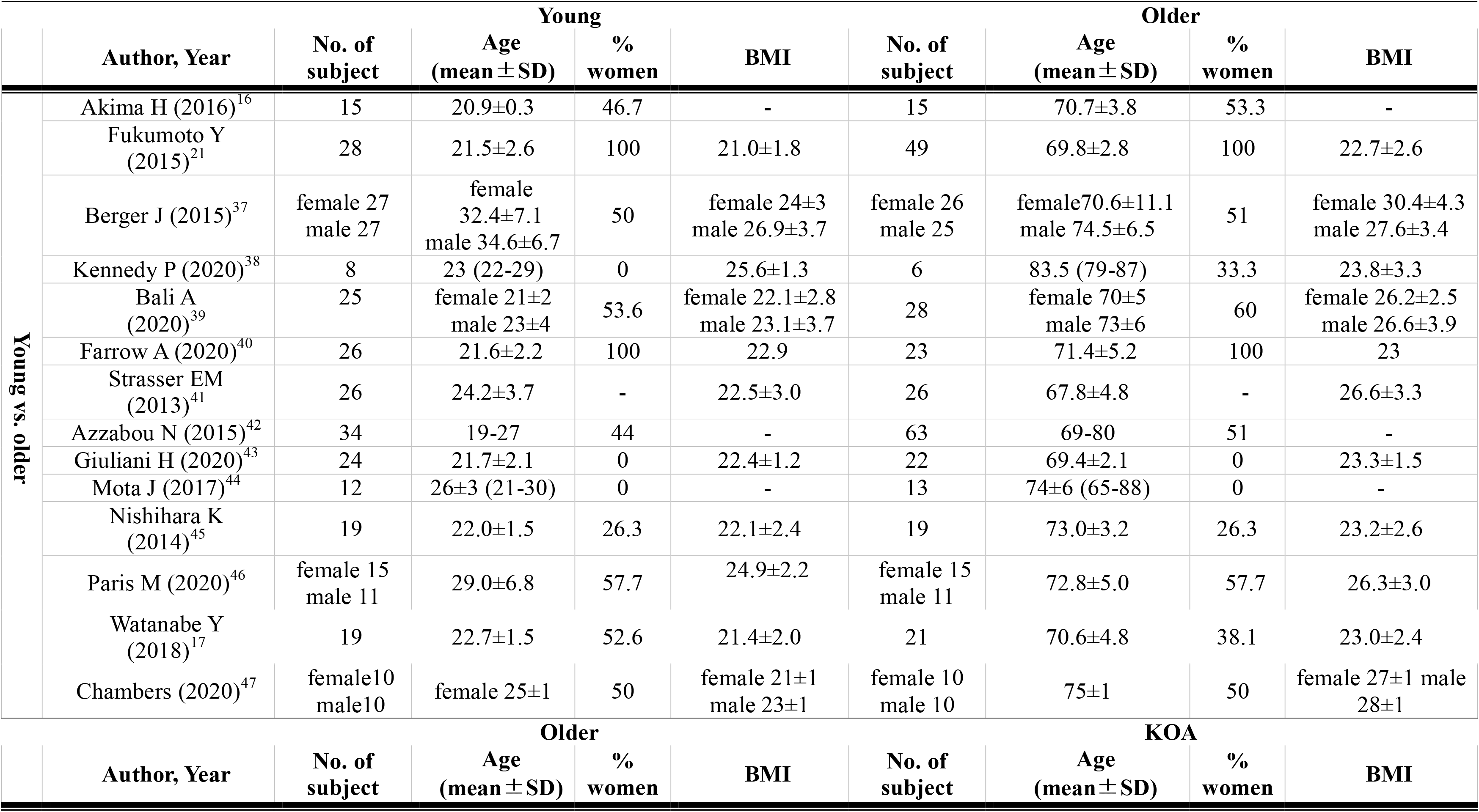
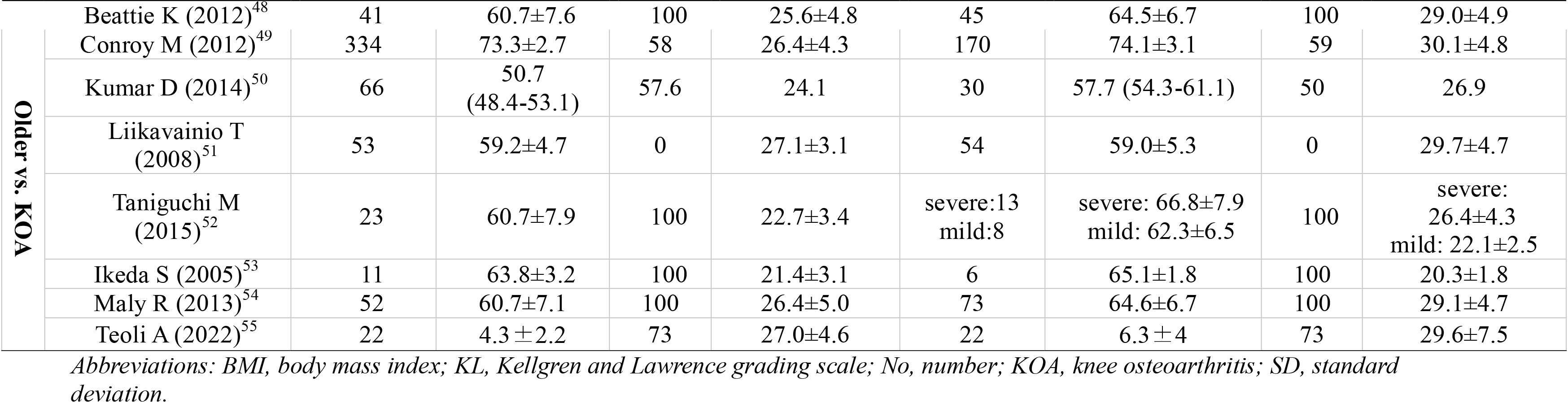
Summary of study participants in the included articles

### US identified the age- and KOA-related decline in quadriceps muscle quality in a manner comparable to MRI and CT

We first sought to assess change in muscle quality over the course of natural aging. A meta-analysis of 18 studies (MRI/CT: 7 studies, US: 11 studies) revealed that older adults displayed poor quadriceps quality when compared to young counterparts (**Figure 1A**). When studies reported results separately according to participants’ characteristics (e.g., sex) and disease severity (e.g., KL grade), data were pooled separately in the analysis to consider their potential confounding effects on fat deposition in skeletal muscle. When comparing across the different imaging modalities, we observed no difference between gold standard methods (i.e., MRI/CT) versus US in the identification of altered quadriceps muscle quality (*p* = 0.41, *I^2^* = 0%), indicating that the age-related decline in muscle quality can be reliably detected by US (**Figure 1A**). Inter-study heterogeneity was high in the US studies (*I^2^* = 69%, *p*=0.0003). Meta-regression and subgroup analyses identified no variables that could significantly explain the high extent of heterogeneity in the age-related decline in quadriceps muscle quality (**Table S4-S5**). However, leave-one-out analysis revealed that the high heterogeneity, defined as *I*^2^ >50%, was significantly improved (from 69% to 35%) by omitting one study^21^ (**Table S6**). This finding suggests that this study was largely responsible for the high inter-study heterogeneity in our data. Nevertheless, participants’ characteristics and disease severity in this study^21^ were similar to the other included studies, suggesting the existence of unknown confounders.

**Figure 1.**
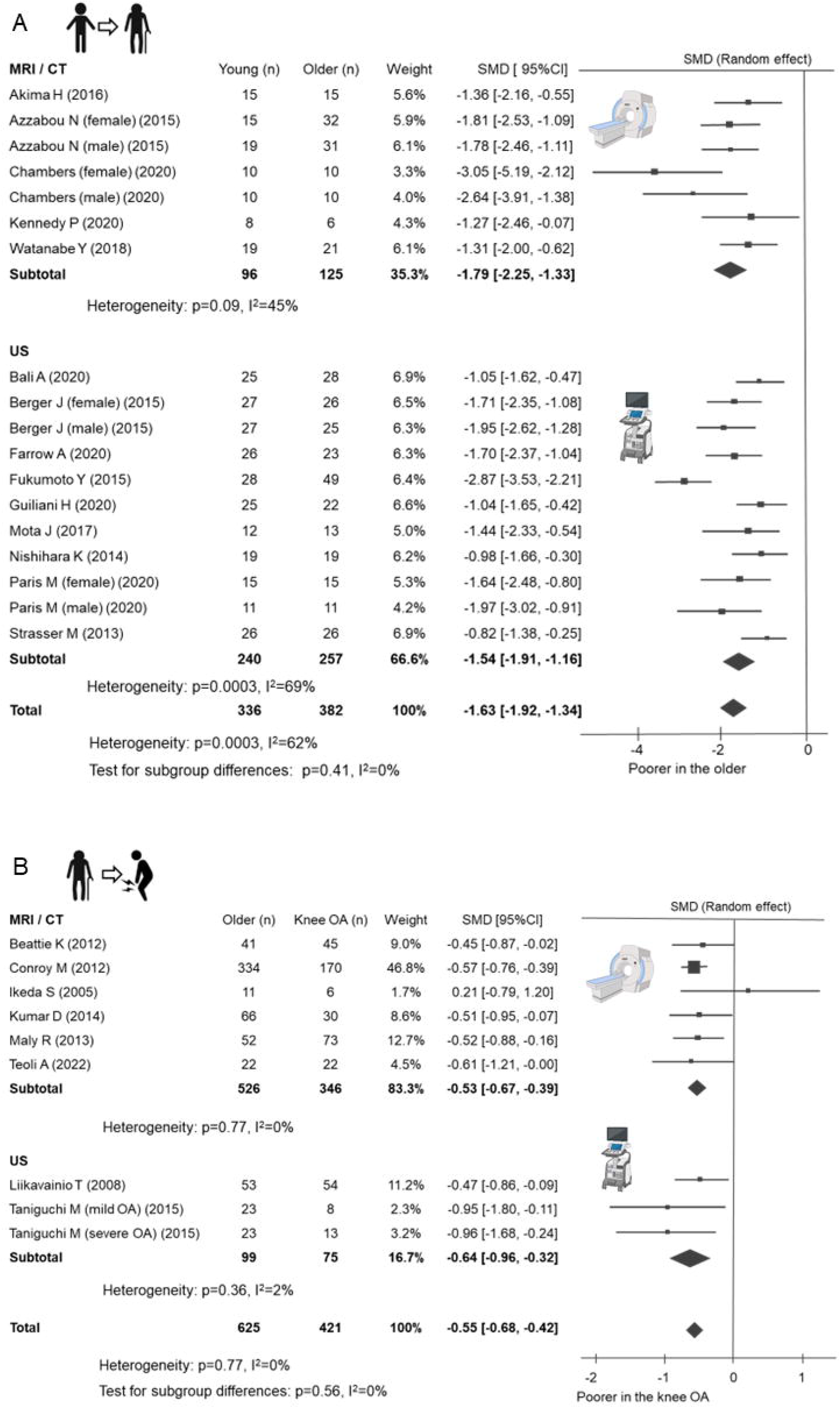
Forest plot of subgroup analysis in measuring devices for young vs older (A) and older vs. KOA (B) The black diamond represents the pooled effect size. The vertical solid line at 0 represents no difference. *Abbreviations: 95%CI, 95% confidence interval; CT, computed tomography; KOA, knee osteoarthritis; MRI, magnetic resonance imaging; SMD, standard mean difference; US, ultrasonography*.

We next performed a meta-analysis of the 9 available studies that included healthy individuals versus individuals with KOA (MRI/CT: 6 studies, US: 3 studies) in order to determine whether osteoarthritic pathological processes aggravate the age-induced decline in quadriceps muscle quality. As expected, people with KOA displayed poorer quadriceps muscle quality compared to age-matched older adults (**Figure 1B**). Similar to the observed age-related decline in quadriceps muscle quality, US had a discriminative ability to identify OA-related decline in quadriceps muscle quality that was comparable to MRI and CT (*p* = 0.56, *I^2^* = 0%) (**Figure 1B**).

Since US offers advantages in clinical practice, including improved cost-efficiency and accessibility, we sought to determine whether US assessment for the aging- and KOA-related altered quadriceps muscle quality was reliable according to the quality of evidence by the Grades of Recommendation, Assessment, Development, and Evaluation (GRADE) system. The GRADE determined that publication bias is possible in the US-based comparison of older vs. KOA due to the small number of studies included (i.e., <10 studies; **Table 2**). As such, more research is needed to characterize KOA-related alterations in quadriceps muscle quality in order to establish US as a reliable means of identifying adults at high risk of KOA in the clinic.

**Table 2.**
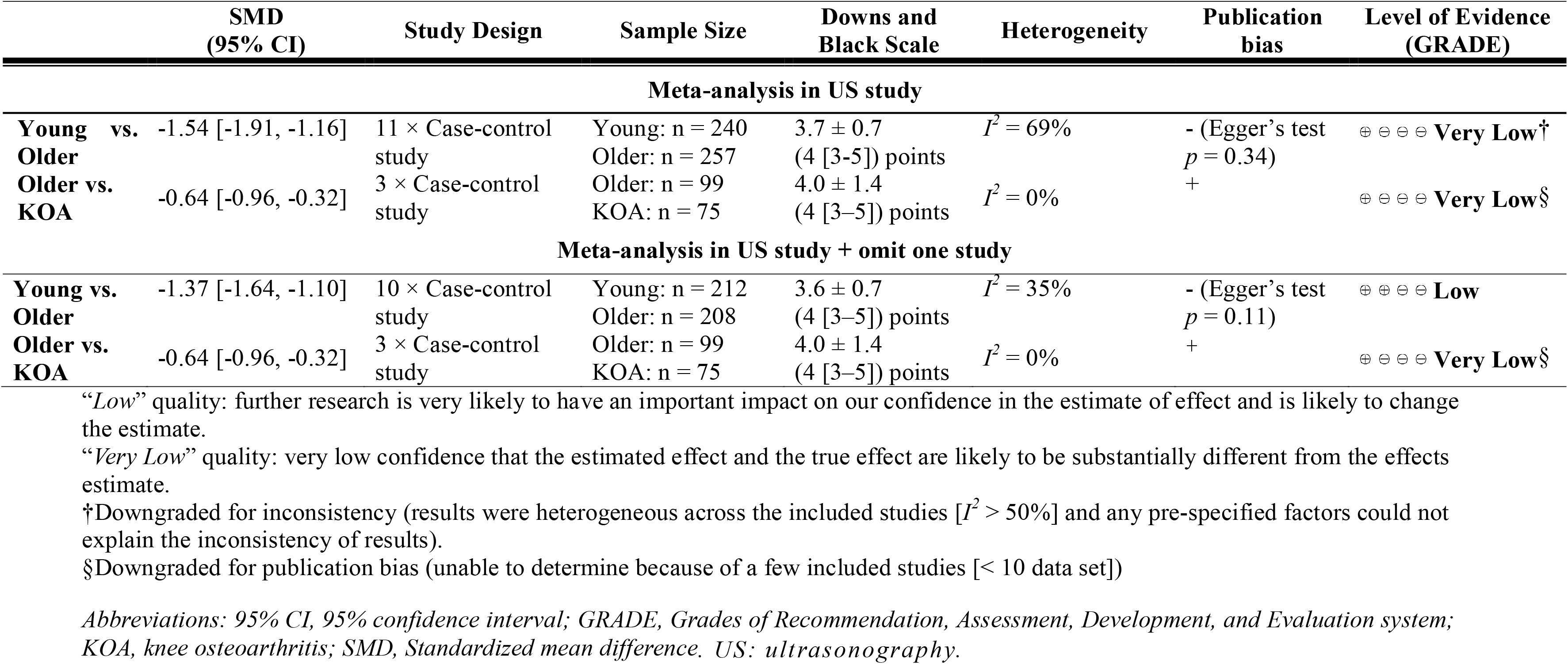
Summary of the body of evidence according to the GRADE’s approach

### Fat deposition in vastus medialis muscle predicted both the onset and development of KOA

To more comprehensively describe the relationship between age- and KOA-related decline in quadriceps muscle quality, we performed a network meta-analysis. Network meta-analysis allows for analysis of the results across multiple independent studies. This analysis method utilizes evidence from direct comparisons (e.g., trials directly comparing treatment A and B) and indirect comparisons (e.g., the combination of trials comparing A with C and trials comparing B with C).^22^ In this study, muscle quality in young adults was set as the control group so that network meta-analysis can be used to directly compare the decline in muscle quality with aging (i.e., healthy young vs healthy older) and in the setting of KOA (i.e., healthy young vs. KOA). Results revealed that the decline in muscle quality in the setting of KOA (SMD:2.07, 95% CI:1.86, 2.27) is significantly faster than with healthy aging (SMD:1.53, 95% CI:1.39, 1.66). This finding supports our meta-analysis demonstrating that the decline in quadriceps muscle quality seen during healthy aging is aggravated by osteoarthritic pathological changes in the knee (**Figure 2A**, **2B**).

**Figure 2.**
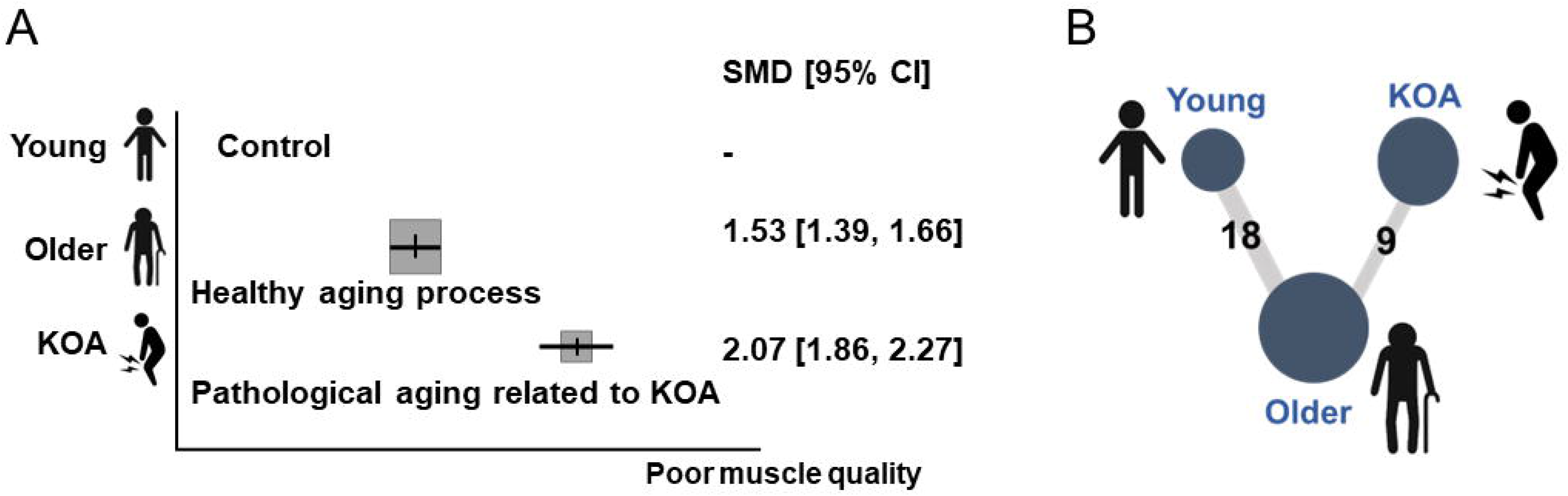
Network meta-analysis comparing healthy aging and pathological aging related to KOA **A,** A decline in quadriceps muscle quality during pathological aging related to KOA was greater than healthy aging. Healthy young adults were set as controls. **B,** Network diagram showing the comparison of number of participants and studies. Each node represents the group (young, older, and KOA). Size of node and edge represents the number of participants and studies, respectively (young vs. older, 18 studies with 718 participants; older vs. KOA, 9 studies with 1,046 participants). *Abbreviations: 95% CI, 95% confidence interval; KOA, knee osteoarthritis; SMD, standard mean difference;*.

Finally, to validate and translate the findings of the network meta-analysis to a clinical setting, we performed a case-control trial of muscle quality assessments using US. The case-control trial was designed to address two key questions that could not be assessed by the meta-analysis: 1) *which of the target quadriceps muscles should be selected for analysis*, and 2) *does the degree of fatty infiltration in skeletal muscle vary with disease severity?*

Based on the calculated required sample size (see Supplemental method), this trial initially recruited 28 participants. According to pre-determined exclusion criteria (Supplemental method), of the 28 participants, three (10.7%) healthy young or older adults were excluded due to intense low back pain (one healthy young adult) and the presence of knee symptoms (two healthy older adults). Thus, 25 participants that met pre-defined inclusion criteria were ultimately included (healthy young: n = 8, average age: 25.9 years, 63% women; healthy older: n = 8, average age: 69 years, 75% women; individuals with KOA: n = 8, average age: 69.1 years, 67% women; see **Table S7** for details). A generalized linear mixed model revealed that corrected echo intensity of VM was lowest (i.e., greatest muscle quality) in the healthy young individuals, followed by healthy older individuals, and then lastly in individuals with KOA (*p*=0.001, 95% CI: 7.42, 24.07). On the other hand, this relationship was not significantly observed in the RF (*p*=0.193, 95% CI: -3.56, 18.56). With the US measurement setting used in the current study (see Supplementary Methods), a decline in muscle quality with aging and in the setting of pathology related to KOA (7.3 a.u. [95% CI: 6.6, 8.1] per 10 years) was significantly greater than that observed with healthy aging (4.6 a.u. [95% CI: 3.8, 5.3] per 10 years) (**Figure 3A**). This correlation was consistent with the network meta-analysis shown above.

**Figure 3.**
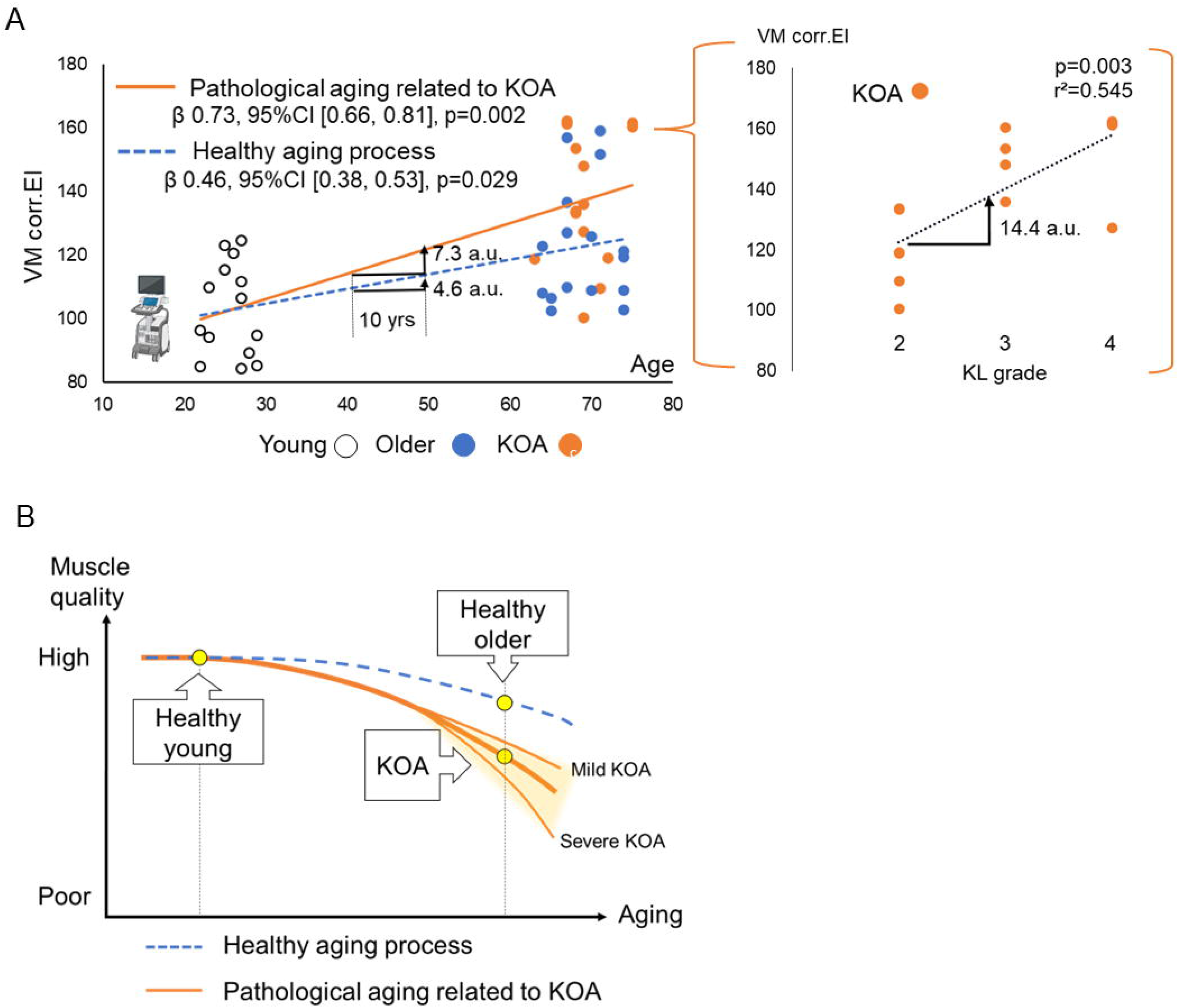
Validation case-control study characterizing quadriceps muscle quality alterations during healthy aging versus pathological aging relating to KOA as assessed by ultrasonography **A,** Scatter plot of VM corrected EI in healthy young (8 participants, 14 knees), healthy older (8 participants, 16 knees), or people with KOA (9 participants, 14knees). Knees with a history of injury were excluded from analysis. Corrected EI of VM increased in healthy aging process (β: 0.73, p=0.002) and pathological aging related to KOA (β: 0.46, p=0.029), which was much higher with increasing KL grade. Regression line and p-value were calculated by generalized linear mixed model with age as a dependent variable. Under the conditions of the present measurement, corrected EI of VM increased at a rate of 4.6 a.u. per 10 years in healthy aging process and 7.3 a.u. per 10 years in pathological aging related to KOA. **B,** Graphical abstract summarizing the findings of the proof-of-concept trial. Fatty infiltration in VM muscle with natural aging process is accelerated with KOA. These trajectory differences may be clinically assessable by ultrasonography, a useful approach to identify the adult with high risk of KOA. *Abbreviations: 95% CI, 95% confidence interval; a.u., arbitrary unit;* β*, regression coefficient; Corr.EI, corrected echo intensity; KOA, knee osteoarthritis; VM, vastus medialis; yrs, years*.

To gain further insight into the decline in muscle quality associated with KOA, we performed a subgroup analysis of our case-control study that aimed to determine whether individuals with an increased severity of KOA displayed a greater decline in quadriceps muscle quality. Among the sub-population of KOA, individuals with greater disease severity assessed by KL grade displayed poorer quality in the VM muscle (14.4 a.u. [95% CI: 8.12, 21.85] per additional KL grade) (**Figure 3A**), a value greater than 10 years change in the natural aging process. Taken together, these studies suggested that VM muscle quality, which is a suitable target in assessing KOA, not only decline in natural aging process, but accelerate with the onset as well as development of KOA (**Figure 3B**).

## DISCUSSION

This systematic review with network meta-analysis showed that KOA exacerbates the age-related decline in quadriceps muscle quality over time. Notably, a series of subgroup analyses indicated that the decline in muscle quality was adequately identified by US. Further, US outcomes were largely comparable to MRI or CT, two gold standard methods to assess muscle quality. To our knowledge, this is the first comprehensive meta-analysis summarizing the trajectory in muscle quality change during healthy aging and in the setting of KOA (**Figure 3B**). The findings from the meta-analyses were validated by the case-control study. The study also addressed the existing gap uncovered by the meta-analyses by providing the first evidence that VM is the most appropriate quadriceps muscle for US-based assessment of muscle quality over time, and that radiographic disease severity displayed a greater impact on fatty infiltration than aging. In this case-control study, we provided reference values for the qualitative changes in VM muscle associated with natural aging and KOA, as quantified by US.

Here, we focused on fat infiltration as an indicator of the decline in muscle quality.^23,24^ Indeed, inter-and intra-muscular fat has an endocrine role implicated in the pathogenesis of OA via the secretion of adipokines,^6,7^ but the role of adipokines is only one piece of the inflammatory picture of KOA. Once previously thought to be a mechanical disease of wear and tear,^25^ KOA is now recognized as a complex metabolic and inflammatory condition driven by factors including adipokines. In the context of obesity-induced KOA, the impact of obesity extends beyond excessive joint loading, and also involves persistent low-grade inflammation due to adipokines secreted from adipose tissue (e.g., leptin and IL-6). Serum levels of leptin are associated with reduced cartilage thickness,^26^ while serum-levels of IL-6 are associated with joint space narrowing and cartilage loss in older adults.^27^ These processes likely contribute to the critical role of intramuscular fat in the onset and progression of KOA.^7,28^ It is additionally thought that acute swelling, pain, and inflammation in people with KOA may lead to decreased mobility, resulting in fat accumulation within the muscle.^29,30^ This bidirectional interaction between fat infiltration and osteoarthritis changes about the knee is an area of interest for future longitudinal studies.

In the present study, the findings from meta-analyses implicate that US can adequately identify increased fat infiltration of the quadriceps in the clinical setting in a manner comparable to MRI and CT. Intriguingly, the results of the subsequent case-control study suggested that the VM muscle is more sensitive in the identification of decline of muscle quality compared to the RF muscle, which is also frequently targeted in US assessment as indicated by our systematic review. Our case-control study supports previous longitudinal MRI studies demonstrating that fatty infiltration in the VM muscle, but not the other individual quadriceps (RF, VL, and VI), was significantly associated with structural abnormalities in people with KOA, such as cartilage loss and bone marrow lesions.^31^ As such, early detection of VM muscle quality decline may be a valuable tool to identify older adults at high risk or early stages of the disease, given the growing evidence that changes in muscle quality may precede a loss of muscle mass, ultimately leading to functional weakness.^32,33,34^ Our case-control study proposed reference values of VM muscle quality decline assessed by US (**Figure 3A**), which builds a foundation towards establishment of an US-based system to predict adults at risk for KOA progression.

Although this study provides a new perspective for the decline in quadricep muscle quality during healthy and pathological aging related to KOA, it has some limitations. Firstly, in the current meta-analysis, a limited number of studies were included for comparison of KOA and age-matched healthy older adults, thereby decreasing the quality of evidence as assessed by GRADE. Secondly, because the majority of participants were women in the meta-analysis and case-control study, the interpretation may not necessarily be generalizable to males.^35^ Lastly, lower inter/intra-rater reliability of US assessment may limit the application in the clinic, particularly for longitudinal follow-up among different assessors. This study calls for future studies following up the muscle quality changes with standardized US procedure.

Taken together, these studies suggest that US is a promising tool to identify early KOA given its capacity to reliably assess accelerated fatty infiltration in the VM muscle. These findings bring us one step closer towards establishing the way to identify adults at risk for KOA in clinical practice.

## METHODS

### Systematic review, meta-analysis, and network meta-analysis

The full methodology for systematic review with meta-analysis is provided in the supplemental file and includes information on: eligibility criteria, literature search, study selection, data collection, determining inclusion, data extraction, meta-analysis, network meta-analysis, and quality of evidence (GRADE). Briefly, a systematic review identified articles studying age- and KOA-related change in quadriceps muscle quality according to clinically available imaging techniques (i.e., ultrasound, MRI, or CT). Summary results were obtained through pairwise meta-analysis and network meta-analysis. The protocol of this meta-analysis has been registered in PROSPERO (CRD42022380856).

### US-implemented case-control study

The full methodology for case-control study conducted for validation purpose is provided in the supplemental file. Briefly, this study included participants with healthy young, healthy older, and individuals with KOA group, based on required sample size (n = 24) calculated *priori*. Echo intensity of VM and RF muscle, an indicator for fatty infiltration, was assessed with B-mode ultrasound images,^36^ and represented using grayscale corrected by subcutaneous fat thickness.^15^ A generalized linear mixed model was used to assess alterations in quadriceps muscle during healthy aging (young vs. older) and pathological aging related to KOA (young vs. KOA).

## Data Availability

All data produced in the present study are available upon reasonable request to the authors.

## Acknowledgements

This study was supported in part by a JSPS KAKENHI (Grant-in-Aid for Early-Career Scientists; Grant Number: 18045240) from the Japan Society for the Promotion of Science (https://www.jsps.go.jp/). The funders had no role in study design, data collection and analysis, decision to publish, or preparation of the manuscript.

