## Supplementary material for "Ultrasound-detected accelerated fat deposition in the vastus medialis as a marker for early knee osteoarthritis": FigureS1-S2 and TableS1-S8: Supplementary-SR [clean].docx

**Supplementary Online Content**

**A network meta-analysis integrated with clinical study implicates ultrasound-detected accelerated fat deposition as a sensitive marker of early knee osteoarthritis**

Atsushi Hoki, PT^1^, Ella D'amico, MD^2^, Fabrisia Ambrosio, PhD, MPT^3^,

Yoshikazu Matsuda, MD^4,5^, *Hirotaka Iijima, PhD, PT^6,7^

^1^Department of Physical Therapy, Matsuda Orthopedic Clinic, Saitama, Japan

^2^Department of Physical Medicine and Rehabilitation, University of Pittsburgh, Pittsburgh, PA

^3^Department of Physical Medicine and Rehabilitation, Spaulding Rehabilitation Hospital and Harvard Medical School, Boston, MA

^4^Department of Orthopedic Surgery, Matsuda Orthopedic Clinic, Saitama, Japan

^5^Department of Pharmacy, Josai University, Saitama, Japan

^6^Institute for Advanced Research, Nagoya University, Nagoya, Japan

^7^Biomedical and Health Informatics Unit, Department of Integrated Health Science, Graduate School of Medicine, Nagoya University, Nagoya, Japan

**The PDF file includes:**

Figure S1. Review flow diagram

Figure S2. Funnel plot of comparison in young and older (A), older and KOA (B)

Table S1. Summary of outcome in KOA

Table S2. Summary of measurement methods

Table S3. Summary of outcome

Table S4. Meta-regression analysis

Table S5. Subgroup analysis

Table S6. Leave-one-out analysis

Table S7. Subject characteristics

Table S8. Summary of Down and Black scale

Supplemental references

This supplementary material has been provided by the authors to give readers additional information about their work

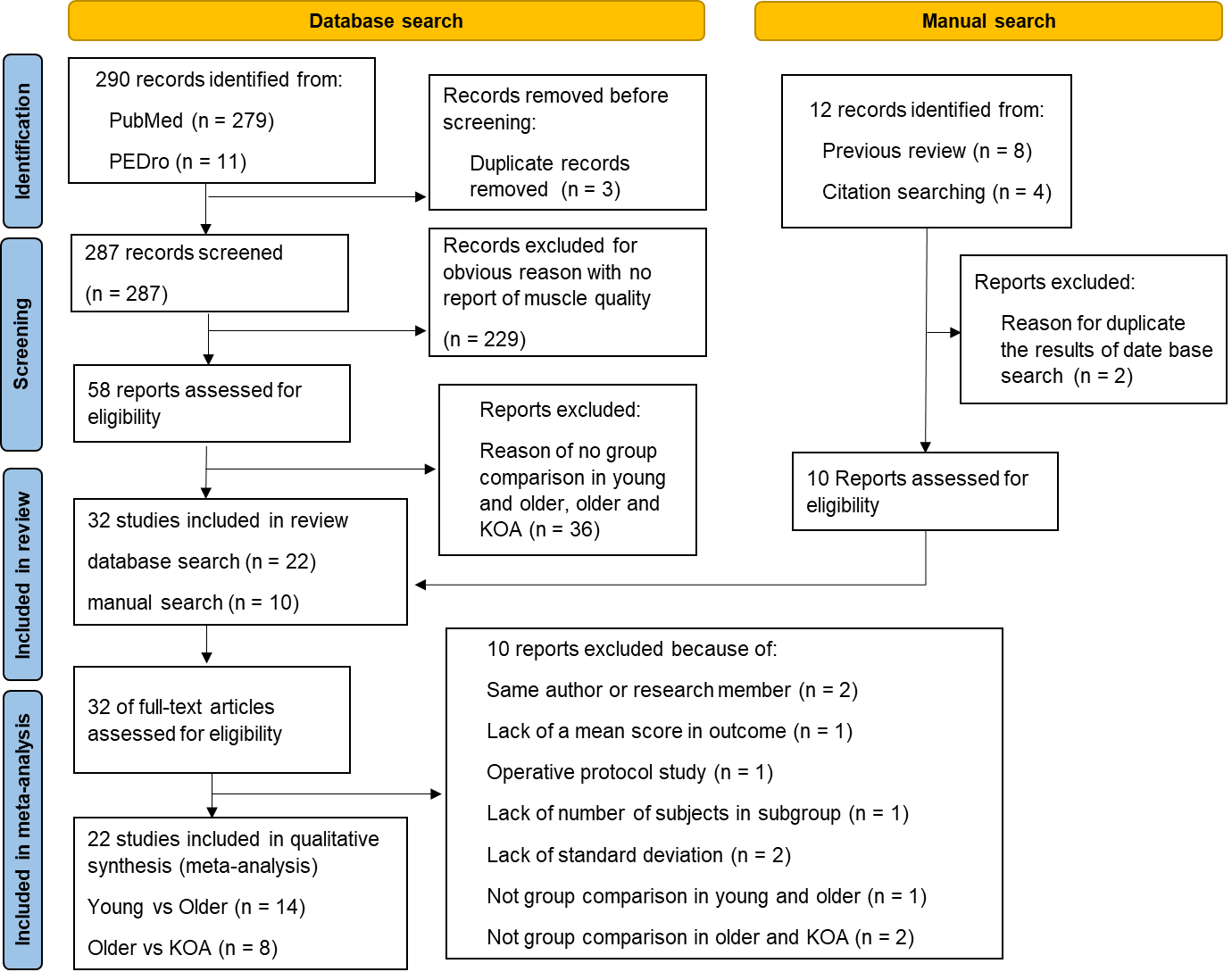

**Figure S1.** Review flow diagram

*Abbreviations: KOA, knee osteoarthritis; n, number.*

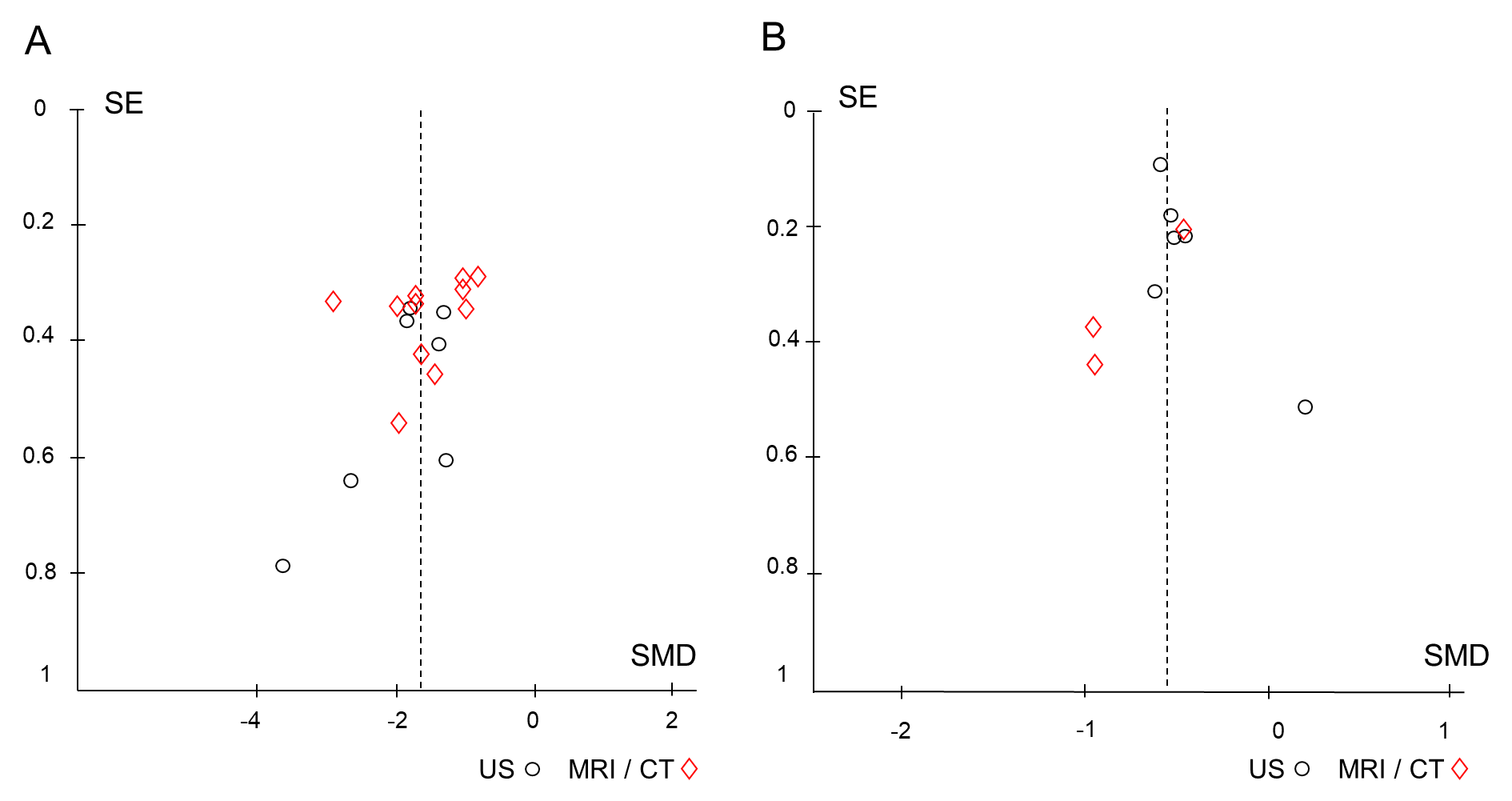

**Figure S2.** Funnel plot of comparison in young and older (A), older and KOA (B)

*Abbreviations : CT, computed tomography; KOA, knee osteoarthritis; MRI; magnetic resonance imaging; SE; standard error, SMD; standardized mean difference; US; ultrasonography.*

**Table S1.** Summary of disease severity and clinical symptom of patients with KOA

| **Author, Year** | **KL grade** | **Clinical symptom** |
| --- | --- | --- |
| Beattie K (2012)^1^ | 2, 3, 4 | 4.2 ± 3.6 (WOMAC, point) |
| Conroy M (2012)^2^ | 2, 3, 4 | - |
| Kumar D (2014)^3^ | 2, 3, 4 | 78.8 (71.4, 86.3) (KOOS pain, point) |
| Liikavainio T (2008)^4^ | 1, 2, 3, 4 | 39.8 (RAND36, point) |
| Taniguchi M (severe OA) (2015)^5^ | 3, 4 | 43.4 ± 26.6 (VAS, mm) |
| Taniguchi M (mild OA) (2015)^5^ | 1, 2 | 15.8 ± 13.5 (VAS, mm) |
| Ikeda S (2005)^6^ | 2 | - |
| Maly R (2013)^7^ | 2, 3, 4 | - |
| Teoli A (2022)^8^ | 1, 2, 3, 4 | Constant pain 17/100  Intermittent pain 31/100  (ICOAP, point) |

*Abbreviations : ICOAP, Intermittent and Constant Osteoarthritis Pain; KL,* *Kellgren and Lawrence grading scale; KOA, knee osteoarthritis; KOOS,* *Knee Injury and Osteoarthritis Outcome Score; RAND36,* *Research and Development 36-Item Health Survey; VAS, visual analogue scale; WOMAC, Western Ontario and McMaster Universities Osteoarthritis Index.*

**Table S2.** Summary of measurement methods

| **Device** | **Outcome** | **Reliability** | **No. of study** |
| --- | --- | --- | --- |
| MRI | Intra muscular fat (cm^2^/ cm^3^/ %) | Intra-later; excellent (0.970)^3^  Inter-later; excellent (0.970)^9^ | 5 (62.5%) |
|  | Inter muscular fat (cm^3^/ %) | Intra-later; excellent (0.950)^7^  Inter-later; excellent (0.950-0.997)^1,7^ | 3 (37.5%) |
| CT | Intra muscular fat (H.U.) | Inter-later; good (0.850)^2^ | 2 (66.7%) |
|  | Inter muscular fat (cm^2^) | - | 1 (33.3%) |
| US | Echo intensity (a.u.)  (Reflect intramuscular fat and fibrous tissue) | Intra-later; moderate to excellent  (0.640 – 0.900)^5,10^  Inter-later; moderate to good  (0.500 – 0.720)^10,11^ | 8 (72.7%) |
|  | Corrcted echo intensity (a.u.)  (Echo intensity that corrected by subcutaneous fat thickness) | Intra-later; excellent (0.950)^12^  Inter-later; good (0.782)^13^ | 3 (27.3%) |

*Abbreviations :* *a.u., arbitrary unit; CT, computed tomography; H.U., Hounsefield Unit; MRI; magnetic resonance imaging; No, number; US, ultrasonography.*

**Table S3.** Summary of outcome

|  | **Author** | **Device** | **Outcome** | **Target muscle** | **Gain*** | **ICC** | **Funding score** |
| --- | --- | --- | --- | --- | --- | --- | --- |
| **Young vs older** | Akima H (2016)^9^ | MRI  US | Intramuscular fat (%) | VL | - | Interlater: 0.970 | 1 |
|  | Fukumoto Y (2015)^14^ | US | Echo intensity (a.u.) | RF + VI | 58 | - | 0 |
|  | Berger J (2015)^10^ | US | Echo intensity (a.u.) | RF | 58 | Intralater: 0.640  Interrlater: 0.500 | 1 |
|  | Kennedy P (2020)^15^ | MRI | Fat fraction (%) | VM | - | - | 1 |
|  | Bali A (2020)^16^ | US | Echo intensity, Corrected echo intensity (a.u.) | VL + RF | 55 | - | 0 |
|  | Farrow A (2020)^12^ | US | Corrected echo intensity (a.u.) | VL | 50 | Intralater: 0.950 | 0 |
|  | Strasser EM (2013)^17^ | US | Echo intensity (a.u.) | VM | - | - | 0 |
|  | Azzabou N (2015)^18^ | MRI | Fat ratio (%) | VM | - | - | 1 |
|  | Giuliani H (2020)^11^ | US | Echo intensity (a.u.) | VM + VL + RF | - | Interlater: 0.720 | 1 |
|  | Mota J (2017)^13^ | US | Corrected echo intensity (a.u.) | RF | 50 | Interlater: 0.782 | 0 |
|  | Nishihara K (2014)^19^ | US | Echo intensity (a.u.) | RF | - | - | 0 |
|  | Paris M (2020)^20^ | US | Echo intensity (a.u.) | RF | - | - | 1 |
|  | Watanabe Y (2018)^21^ | CT  US | H.U. | Total thigh | - | - | 1 |
|  | Chambers (2020)^22^ | MRI | Intermuscular adipose tissue(%) | Total thigh | - | - | 1 |
| **Older vs KOA** | Beattie K (2012)^1^ | MRI | Intermuscular fat volume (cm^3^) | VM + VL + VI + RF | - | Interlater: 0.997 | 1 |
|  | Conroy M (2012)^2^ | CT | H.U., Intermuscular fat (cm^2^) | Total thigh | - | Interlater: 0.850 | 1 |
|  | Kumar D (2014)^3^ | MRI | Intramuscular fat volume (cm^3^) | VM + VL + VI + RF | - | Intralater: 0.970  [95%CI: 0.920-0.990] | 1 |
|  | Liikavainio T (2008)^4^ | US | Echo intensity (a.u.) | RF | - |  | 0 |
|  | Taniguchi M (2015)^5^ | US | Echo intensity (a.u.) | VM | 58 | Intralater: 0.830-0.900 | 0 |
|  | Ikeda S (2005)^6^ | CT | Intermuscular fat (cm^2^) | Total thigh | - | - | 0 |
|  | Maly R (2013)^7^ | MRI | Intermuscular fat volume (cm^3^) | VM + VL + VI + RF | - | Intralater > 0.950  Interlater > 0.950 | 0 |
|  | Teoli A (2022)^8^ | MRI | Intramuscular fat (fat %) | VM | - | - | 1 |

*Gain is a uniform amplification of the ultrasonic signal. Echo intensity has varied by ultrasonography settings represented gain.

*Abbreviations: a.u., arbitrary unit; CT, computed tomography; EI, echo intensity; H.U., Hounsefield Unit; ICC, intraclass correlation coefficients; IMF, intermuscular fat; KOA, knee osteoarthritis; LDMA, low-density muscle area; MRI, magnetic resonance imaging; RF, rectus femoris; US, ultrasonography; VI, vastus intermedius; VL, vastus lateralis; VM, vastus medialis.*

**Table S4.** Meta-regression analysis

| **Young vs. Older in US** | | | | | |
| --- | --- | --- | --- | --- | --- |
| **Variable** | **Coefficient** | **Lower 95%CI** | **Upper 95%CI** | **SE** | ***p*-value** |
| Age. young | -0.032 | -0.12 | 0.059 | 0.040 | 0.45 |
| Age. older | -0.049 | -0.26 | 0.16 | 0.094 | 0.62 |
| %female. young | -0.0043 | -0.013 | 0.0047 | 0.0039 | 0.30 |
| %female. older | -0.0042 | -0.013 | 0.0049 | 0.0039 | 0.32 |
| BMI. young | -0.048 | -0.34 | 0.24 | 0.13 | 0.71 |
| BMI. older | 0.017 | -0.19 | 0.22 | 0.089 | 0.85 |
| Year of publication | 0.0025 | -0.15 | 0.16 | 0.069 | 0.97 |
| Down & Black | 0.14 | -0.55 | 0.82 | 0.30 | 0.66 |
| Sample size  (young + older) | -0.013 | -0.040 | 0.014 | 0.012 | 0.29 |
| Ultrasound set up (gain) | -0.074 | -0.27 | 0.12 | 0.069 | 0.34 |

Since the high extent of inter-study heterogeneity was found in the young vs. older studies with US, meta-regression analysis was performed for each of continuous variables.

*Abbreviations: 95% CI, 95% confidence interval; BMI, body mass index; SE, standard error.*

**Table S5.** Subgroup analysis

| **Young vs. Older in US** | | | | | |
| --- | --- | --- | --- | --- | --- |
| **Variable** | **Mean** | **Lower95%CI** | **Upper95%CI** | **SD** | ***p*-value** |
| Target muscle  VM (n=1)  VL (n=1)  RF (n=6)  Quadriceps mean (n=3) | -0.82  -1.53  -1.62  -1.65 | NA  NA  -2.00  -4.27 | NA  NA  -1.23  -0.96 | NA  NA  0.37  1.05 | 0.70 |
| Funding score  yes (n=5)  no (n=6) | -1.66  -1.48 | -2.13  -2.27 | -1.19  -0.68 | 0.38  0.76 | 0.63 |

Subgroup analyses were performed for categorical variables because of the high extent of inter-study heterogeneity found in the young vs. older studies with US.

*Abbreviations: 95% CI, 95% confidence interval; n, number; NA, not applicable; RF, rectus femoris; SD, standard deviation; VM, vastus medialis; VL, vastus lateralis.*

**Table S6.** Leave-one-out analysis

| **Young vs. Older in US** | | | |
| --- | --- | --- | --- |
| **Omitting study** | **US study** | | **Subgroup difference** |
|  | **SMD [95%CI]** | ***I^2^* (%)** | ***p*-value** |
| [None] | -1.54 [-1.91, -1.56] | 69 | 0.41 |
| Bali A^16^ | -1.60 [-2.00, -1.19] | 70 | 0.54 |
| Berger J (female)^10^ | -1.52 [-1.94, -1.11] | 72 | 0.40 |
| Berger J (male)^10^ | -1.50 [-1.90, -1.09] | 71 | 0.35 |
| Farrow A^12^ | -1.52 [-1.94, -1.11] | 72 | 0.40 |
| Fukumoto Y^14^ | -1.37 [-1.64, -1.10] | 35 | 0.13 |
| Guiliani H^11^ | -1.59 [-2.00, -1.19] | 70 | 0.53 |
| Mota J^13^ | -1.55 [-1.95, -1.14] | 72 | 0.44 |
| Nishihara K^19^ | -1.60 [-2.00, -1.20] | 70 | 0.54 |
| Paris M (female)^20^ | -1.53 [-1.94, -1.12] | 72 | 0.41 |
| Paris M (male)^20^ | -1.51 [-1.91, -1.11] | 72 | 0.37 |
| Strasser EM^17^ | -1.62 [-2.00, -1.24] | 66 | 0.58 |

Subgroup difference calculated between US subgroup and MRI and CT subgroup.

*Abbreviations: 95% CI, 95% confidence interval; CT, computed tomography; MRI, magnetic resonance imaging; SMD, standardized mean difference; US, ultrasonography.*

**Table S7.** Subject characteristics of case control study

| **Variables** | **Young** | **Older** | **KOA** |
| --- | --- | --- | --- |
| no. (knees) | 8 (14) | 8 (16) | 9 (14) |
| Age, years | 25.9 ± 2.4 | 69.0 ± 3.9 | 69.1 ± 3.4 |
| %women | 62.5 | 75 | 66.7 |
| BMI, kg/m^2^ | 21.7 ± 2.0 | 22.2 ± 2.9 | 24.6 ± 3.9 |
| KL grade, (no. grade2, 3, 4) | - | - | 6, 4, 4 |
| Analgesic medication, no. (%) | 0 | 1 (12.5) | 1 (11.1) |

*Abbreviations: KOA, knee osteoarthritis; no, number; BMI, body mass index;* *KL grade, Kellgren, and Lawrence grading grade.*

**Table S8.** Summary of Down and Black scale

| **Items in check list** | **Blind**  **(item 15)** | **Data dreading (item 16)** | **Statistical test (item 18)** | **Outcome (item 20)** | | **Time period (item 22)** | **Adjustment (item 25)** | **Total** |
| --- | --- | --- | --- | --- | --- | --- | --- | --- |
| Young vs. Older | | | | | | | |  |
| Akima H (2016)^9^ | 0 | 1 | 1 | 1 | 0 | | 0 | 3 |
| Fukumoto Y (2015)^14^ | 1 | 1 | 1 | 1 | 0 | | 0 | 4 |
| Berger J (2015)^10^ | 0 | 1 | 1 | 1 | 0 | | 0 | 3 |
| Kennedy P (2020)^15^ | 0 | 1 | 1 | 1 | 0 | | 0 | 3 |
| Bali A (2020)^16^ | 0 | 1 | 1 | 1 | 0 | | 1 | 4 |
| Farrow A (2020)^12^ | 0 | 1 | 1 | 1 | 0 | | 1 | 4 |
| Strasser EM (2013)^17^ | 1 | 1 | 1 | 1 | 0 | | 1 | 5 |
| Azzabou N (2015)^18^ | 0 | 1 | 1 | 1 | 0 | | 0 | 3 |
| Giuliani H (2020)^11^ | 0 | 1 | 1 | 1 | 0 | | 1 | 4 |
| Mota J (2017)^13^ | 0 | 1 | 1 | 1 | 0 | | 0 | 3 |
| Nishihara K (2014)^19^ | 0 | 1 | 1 | 1 | 0 | | 0 | 3 |
| Paris M (2020)^20^ | 0 | 1 | 1 | 1 | 0 | | 1 | 4 |
| Watanabe Y (2018)^21^ | 0 | 1 | 1 | 1 | 0 | | 0 | 3 |
| Chambers A (2020)^22^ | 0 | 1 | 1 | 1 | 0 | | 0 | 3 |
| Older vs. KOA | | | | | | | |  |
| Beattie K (2012)^1^ | 1 | 1 | 1 | 1 | 0 | | 1 | 5 |
| Conroy M (2012)^2^ | 0 | 1 | 1 | 1 | 0 | | 1 | 4 |
| Kumar D (2014)^3^ | 1 | 1 | 1 | 1 | 0 | | 1 | 5 |
| Liikavainio T (2008)^4^ | 0 | 1 | 1 | 1 | 0 | | 0 | 3 |
| Taniguchi M (2015)^5^ | 1 | 1 | 1 | 1 | 0 | | 1 | 5 |
| Ikeda S (2005)^6^ | 0 | 1 | 0 | 1 | 0 | | 1 | 3 |
| Maly R (2013)^7^ | 0 | 1 | 1 | 1 | 0 | | 1 | 4 |
| Teoli A (2022)^8^ | 0 | 1 | 1 | 1 | 1 | | 0 | 4 |

*Abbreviations: KOA, knee osteoarthritis.*

**SUPPLEMENTARY METHODS**

***Eligibility criteria, literature search, and study selection***

Systematic review with meta-analysis was conducted in accordance with the Preferred Reporting Items for Systematic reviews and Meta-Analyses (PRISMA) statement^23^ PRISMA protocols (PRISMA-P),^24^ Meta-analysis of Observational Studies in Epidemiology (MOOSE) checklist,^25^ PRISMA for Network Meta-Analysis (PRISMA-NMA),^26^ and the Cochrane Handbook for Systematic Reviews of Interventions.^27^

PubMed and Physiotherapy Evidence Database (PEDro) electronic databases were used for this work. Searches in the PubMed used combined key terms, including “Aged,” “Skeletal muscle,” “Quadriceps,” “Ultrasonography,” “Computed Tomography, X-ray,” and “Magnetic Resonance Imaging,” using Medical Subject Headings terms. Search in the PEDro used combined key terms, including “Quadriceps muscle, Ultrasonography,” “Quadriceps muscle, Computed Tomography, X-ray” and “Quadriceps muscle, Magnetic Resonance Imaging”. Google Scholar was also used as a complementary search engine. In addition, a manual search of the reference lists of past systematic review^28^ was performed. These citation indices are recommended by the Cochrane Handbook.^27^

Included studies met the following pre-defined criteria: (1) published in a peer review journal, (2) written in English, (3) participants with healthy young and healthy older and/or those with healthy older and people with KOA, and (4) outcomes included metrics of quadriceps muscle composition (e.g. inter- and intra-muscular fatty infiltration)^29^ assessed by clinically available imaging techniques (i.e., ultrasound, MRI, or CT) . Although some studies defined muscle quality as peak torque per muscle area (e.g. muscle torque, muscle activation), we excluded these studies because these outcome variables would be strongly affected by muscle fatigue and pain, common characteristics in people with KOA.^30,31^ Studies with participants who had undergone total joint arthroplasty, rheumatoid arthritis and history of knee fracture were also excluded. In this review, KOA was defined either radiographically or clinically by using some established existing criteria for OA, such as the American College of Rheumatology criteria.^32^ To maximize sample size in the meta-analysis, this study did not set a strict definition for age in young and older adults and the definition followed individual studies’ criteria. No restrictions were imposed on study dates, follow-up duration, disease severity, involved compartment, or lower limb alignment. The search results were downloaded and filtered in Mendeley Desktop (ver1.19.8). For each electronic database, the end point was March 23 in 2022.

***Determining inclusion and data extraction***

Two content and statistical expert reviewers (AH, ED) assessed eligibility in accordance with the Cochrane Handbook.^27^ The reviewers screened the title and abstracts yielded by the search. The full manuscripts of the articles that met the eligibility criteria were then obtained and reviewed. During these processes, the reviewers prepared and used simple predesigned Google spreadsheets to assess eligibility by extracting study features.

The same reviewers collected the data using a standardized data extraction form including authors, years, subject population, subject age, radiographic disease severity, measuring device, outcome measures, target muscle, intra-rater and inter-rater reliability assessed by intraclass correlation coefficients (ICC). When there was missing or unclear data, authors were contacted. When an article reported muscle quality in quadriceps separately (e.g., vastus medialis and rectus femoris), this study prioritized vastus medialis muscle since longitudinal studies reported that adiposity in vastus medialis is associated with KOA progression.^33,34^

***Data synthesis and network-meta-analysis***

For the meta-analysis, pooled estimates and 95% CIs for standardized mean differences (SMDs) were calculated using the DerSimonian-Laird method.^35^ The SMDs were calculated for paired samples by using the mean between-group difference (healthy young vs. healthy older, healthy older vs. KOA) divided by the pooled standard deviation (SD). The meta-analyses were performed using Review Manager (RevMan) Version 5.4 (Nordic Cochrane Centre, Cochrane Collaboration, Copenhagen, Denmark). We used a forest plot to represent the meta-analysis results in accordance with a previous study.^36^ The size of the SMD was interpreted using Cohen’s d^37^ (<0.5: small effect size, 0.5–0.8: moderate effect size, and ≥0.8: large effect size). Study heterogeneity, defined as the inter-trial variation in study outcomes, was assessed using *I*^2^, which is the proportion of the total variance explained by inter-trial heterogeneity. If there were outcomes with high heterogeneity, leave-one-out analysis was performed by excluding one study at each meta-analysis. For comparison of muscle quality in young, older, and KOA, we performed network meta-analysis. Network meta-analysis is a technique for comparing three or more interventions simultaneously in a single analysis by combining both direct and indirect evidence across a network of studies.^38^ All statistical analyses were performed using EZR, version 1.54 (Saitama Medical Center, Jichi Medical University, Japan), which is a graphical user interface for R, version 4.0.3 (The R Foundation for Statistical Computing, Austria), a modified version of R commander designed to add statistical functions frequently used in biostatistics.^39^ Statistical significance was set at p < 0.05.

If *I*^2^ was ≥50%, a univariate, random-effects meta-regression was performed using certain parameters selected a priori as follows: (1) age (per year), (2) % female (per percent), (3) body mass index (per kg/m^2^), (4) year of publication (per year), (5) Downs and Black scale score (per point), (6) study sample size (per subject), and (7) ultrasound set up (per gain). These factors were chosen because of their potential association with effects estimate. In the case of measurement by ultrasonography, gain value was added using regression analysis as variability because the echo intensity varied depending on device setting.^20^ We conducted a subgroup analysis for assess differences in (1) measuring device (MRI, CT, and ultrasound), (2) target muscle (vastus medialis, vastus lateralis, rectus femoris, overall quadriceps), and (3) funding source (no/yes). Funding source has been added to subgroup analysis to consider the possible effect on heterogeneity (i.e. industry-sponsored research tends to draw pro-industry conclusions).^25,40^

***Overall quality of evidence***

*Risk of bias assessment*

The same reviewers evaluated the risk of bias of each study by using the Downs and Black scale^41^ that was modified to assess internal validity (minimum: 0 point, maximum: 6 points) in accordance with previous meta-analysis.^42^ All items were scored ‘1’ for fulfilling the criterion or ‘0’ if the criterion was not filled. Publications that did not provide sufficient details to fulfill the criterion were also given a ‘0’ for ‘unable to be determined’. A predefined criterion with 1 point suggests that this criterion has a low risk of bias. This scale is a useful tool for assessing the risk of bias in observational studies^43^ and the methodological quality of both randomized controlled trial (RCT) and non-RCT of treatment.^27^

*Publication bias*

To test for publication bias, we used a funnel plot. We also performed Egger’s test^44^ to test funnel plot asymmetry in comparison between young and older. When comparing older and KOA, since relatively few studies (<10 studies) were found for each outcome and the power of the test was too low to distinguish chance from real asymmetry,^27^ we did not perform Egger’s test.

*Summary of evidence: GRADE approach*

To evaluate the quality of evidence, we used the Grading of Recommendations Assessment, Development and Evaluation (GRADE) approach.^45^ The same reviewers graded the quality of evidence for each primary outcome based on five domains: risk of bias, inconsistency, indirectness, imprecision, and publication bias. Evidence quality was downgraded if (1) no attempts were made to blind the assessors measuring main outcomes; (2) the heterogeneity between trials (*I*^2^) was higher than 50%; and (3) 95% CI of SMD crossed the clinical decision threshold, which was defined as zero in this study. Indirectness was not downgraded because all the studies compared groups outcome directly.

***Case-control study of ultrasound assessment for the validation purpose***

*Participants*

We selected participants of three groups, namely healthy young aged from 20 to 30 years, healthy older and patients with KOA aged from 60 to 75 years. They are recruited at Matsuda orthopedic clinic in Japan, from July 2022 to October 2022. Clinic staffs and outpatients with symptom other than knee were included in healthy young adult and healthy older, respectively.

The following basic eligibility criteria were applied to all the participants: (1) no history of surgery to the extremities, (2) ability to walk without an assistive device, (3) no cognitive dysfunction; (4) no neuromuscular disease; and (5) no trauma to lower extremity. In addition to the basic eligibility criteria, the following criteria was further added to heathy young and healthy older adults: (1) no knee pain and symptom in past one week, as defined by the Knee Injury and Osteoarthritis Outcome Score (KOOS)^46^ is zero. For patients with KOA, the following criteria was further added: (1) Kellgren and Lawrence grading scale^47^ (KL grade) 2-4, (2) symptom on the bilateral or unilateral knee in past one week (KOOS pain and/or KOOS ADL is over zero) (3) no osteonecrosis of the medial femoral condyle, all of which were assessed based on information in medical records. Eligibility criteria was set based on subject characteristics from systematic review. This case-control study was approved by the Ethics Committee of the Medical Corporation Nagomi (No. 20221025). All participants provided signed informed consent.

*Ultrasound settings*

Quality of vastus medialis muscle was assessed by the ultrasound in accordance with an established procedure.^48^ Ultrasound images were acquired using a B-mode imaging device (SNiBLE, Konica Minolta Ltd., Japan) equipped using an 8-MHz linear-array probe. The ultrasound settings were kept consistent across all participants (frequency: 9 MHz, gain: 25 dB, dynamic range: 60). The participants lie down with supine position, and the arms and legs relaxed. Vastus medialis (VM) was determined at the 5cm medial side 20% distal between anterior superior iliac spine and proximal end of patella, and rectus femoris (RF) was determined at the midpoint of anterior superior iliac spine and proximal end of patella.^5^ The head of the probe was maintained perpendicular to the muscles examined. A large amount of water-soluble transmission gel (PROJELLY, Jex Inc., Japan) was applied to the skin to enhance acoustic coupling. The captured ultrasound images were used for the assessment of EI using ImageJ software (version 1.53, National Institutes of Health, USA).^49^ Regions of interest were selected at depths of 1.0-4.0 cm. The EI of the analysis region was represented using grayscale values from 0 (black) to 255 (white), with a higher mean pixel intensity value indicating non-lean tissue and, thus, poor muscle quality. For each participant, three images were taken, and mean value of EI calculated from those. All ultrasound imaging was performed by a well-trained investigator (AH). This investigator's reliability has been previous reported.^50^

To account for the effect of subcutaneous fat thickness on echo measurements, a correction factor was applied to echo measurements (Equation 1)^51^ as recommended in several studies.^52^ ^53^ Subcutaneous fat thickness was measured from the superior border of the superficial aponeurosis to the inferior border of the dermis layer. The formula for the corrected EI was as follows:

*y*_2_ = *y*_1_+ (*x***cf*)	(Equation 1)

*y*_2_, corrected echo intensity; *y*_1_, raw echo intensity; *x*, subcutaneous fat thickness; *cf*, correction factor of 40.5278.

*Statistical analysis of validation study*

The sample size was calculated from pilot data of young adult, healthy older and KOA patients (n = 4/group; n = 12 in total) to detect a significant relationship between group difference and corrected EI of VM muscle in a linear regression analysis using Power and Sample Size Program, version 3.1.6 (Vanderbilt University Medical Center, USA).^54^ Preliminary data indicated that we needed to study 24 subjects to reject the null hypothesis with a probability (power 0.8, type I error 0.05) when groups (1: healthy young, 2: healthy older, 3: KOA) was regressed against corrected EI. After considering the potential 10% dropout rate due to the exclusion criteria and missing data, the required sample size of this study was 26 participants.

The muscle quality differences in three groups (1: young, 2: older, 3: KOA) were evaluated by using generalized linear mixed model to account for the similarity between the right and left extremity. We also examined the extent of changes from young to older (i.e. “healthy aging”), and from young to KOA (i.e. “pathological aging”) by including age (continuous) to generalized linear mixed model as a fixed effect. Additionally, the difference of EI according to OA severity (KL grade) was also assessed by generalized linear mixed model. The analysis was performed using EZR, version 1.54 (Saitama Medical Center, Jichi Medical University, Japan), which is a graphical user interface for R, version 4.0.3 (The R Foundation for Statistical Computing, Austria), a modified version of R commander designed to add statistical functions frequently used in biostatistics. Statistical significance was set at p <0.05.
